# Predicting outcomes of cross-sex hormone therapy in gender dysphoria based on pre-therapy resting-state brain connectivity

**DOI:** 10.1101/2020.07.11.20147538

**Authors:** Teena D Moody, Jamie D Feusner, Nicco Reggente, Jonathan Vanhoecke, Mats Holmberg, Amirhossein Manzouri, Behzad Sorouri Khorashad, Ivanka Savic

## Abstract

Individuals with gender dysphoria experience serious distress due to incongruence between their gender identity and birth-assigned sex. Sociological, cultural, interpersonal, and biological factors are likely contributory, and for some individuals medical treatment such as cross-hormone therapy and gender affirming surgery can be helpful. Cross-hormone therapy can be effective for reducing body incongruence, but responses vary, and there is no reliable way to predict therapeutic outcomes. We used clinical and MRI data before cross-sex hormone therapy as features to train a machine learning model to predict individuals’ post-therapy body congruence (the degree to which photos of their bodies match their self-identities). Twenty-five trans women and trans men with gender dysphoria participated. The model significantly predicted post-therapy body congruence, with the highest predictive features coming from the fronto-parietal and cingulo-opercular networks. This study provides evidence that hormone therapy efficacy can be predicted from information collected before therapy and that patterns of functional brain connectivity may provide insights into body-brain effects of hormones, affecting one’s sense of body congruence. Results could help identify the need for personalized therapies in individuals predicted to have low body-self congruence after standard therapy.

## 1. Introduction

Awareness of gender dysphoria (GD) and of gender incongruence has climbed sharply in recent years. Issues related to self-identity, body image, and medical interventions in GD are challenges for the 21st century, particularly given the high suicide risk [1–4] associated with the disorder. GD, in DSM-5 [5], or gender incongruence in ICD-11 [6], refers to significant distress and/or impairment due to a feeling of incongruence between a person’s experienced gender and their birth-assigned sex. A subset of those who identify as transgender suffer from GD [7], though not all GD experiencing people identify as transgender. GD is often treated with cross-sex hormone therapy [8,9], in many cases followed by gender-affirming surgery [10]. In a large proportion of individuals, these therapies reduce the sense of incongruence and associated dysphoria, as well as ratings of depression, suicide risk, and quality of life [8,9,11,12]. In a non-trivial proportion, however, therapy is unsuccessful as evidenced by the high heterogeneity of quality of life outcomes across studies [11,12], as well as variable improvements in body image [13]. This may be due to inter-individual biological variability, contributions from sociological, cultural, or social factors, or (most likely) a combination of these factors.

The medical profession currently lacks the ability to determine who will respond well to therapy and who will not - a critical piece in moving towards personalized, evidence-based medicine to optimize clinical outcomes and efficacy of therapy. As well, this needs to be considered in the context of rapidly changing societal notions of gender identity. Overall there have been few studies that have examined predictors of clinical outcomes for cross-sex hormone or gender-affirming surgical treatments. A recent study of hormone and surgical treatments [13] found that that body dissatisfaction pre-therapy predicted body dissatisfaction post-therapy (p < 0.001), but there was no predictive value of birth-assigned sex (p = 0.83), age (p = 0.23), or physical “passing” at admission (p = 0.50). A study of surgical treatments found better outcomes were predicted by pre-treatment lower dissatisfaction with secondary sex characteristics (p < 0.001) and less psychopathology (p < 0.028), as well as being homosexual (p < 0.002) (defined in relationship to birth-assigned sex) [14].

In those with GD, the primary therapy outcomes - improving congruence between one’s gender identity and the sex-related physical characteristics of their body, as well as associated dysphoria - are conscious experiences and have a prominent basis in gender roles, psychological and sociological structures, and in neural function. The current study is specifically focused on constructing a preliminary prediction model of cross-hormone therapy response.

We recently proposed a hypothesis that GD is characterized by a functional disconnection between systems in the brain that process the perception of self (“self-referential”) and those that mediate own-body perception [15],[16]. Self-referential systems include the default mode network, particularly medial prefrontal cortical regions such as the dorsal and pregenual anterior cingulate cortex [17], and the salience network, particularly the insular cortex [18], [19]. Involved in own-body perception are the temporoparietal junction [20] and the extrastriate and fusiform body areas [21]. We proposed that differences in coordinated activation and connections between own-body and self-perception networks could explain the discomfort with their bodies reported by individuals with GD [15],[16]. To better understand relationships between body perception and gender-related self-identity, we previously designed a “body morph task” [22], specifically to test the degree of incongruence between self own-body perception in relation to the body sex characteristics. In this task, binary-identifying participants (transgender as well as cisgender individuals) view their own bodies in unitards in photographs that are incrementally morphed with others’ bodies that are the same as, or opposite to, their birth-assigned sex. For each image presentation, the participant assesses to what degree the image represents him/her. We then determine the ratings of the “body morph index”: the perception of the degree of self, represented by an index calculated from ratings across *all* of the morphed bodies presented. The body morph index provides an indication of an individual’s maximal perception of ‘self’ on a continuum from traditionally feminine to traditionally masculine appearances.

Studies using the body morph task [15,22–25] suggest that, at least in trans men, information from this task in combination with resting state fMRI may be an indication of self-perception pre-therapy, as well as the effects of hormone therapy on self-perception. Further, because the body index is calculated from multiple presentations of morphs toward and away from birth-assigned sex, it may be a more objective and precise instrument to assess body congruence compared with a subjective, self-report rating scale, e.g. the Transgender Congruence Scale (TCS), or qualitative reports. Therefore, in the current study we used the body index as our main outcome variable to quantify an individual’s body congruence.

In alignment with our previous studies of structural and functional brain systems in GD, we focused on seven brain networks as potential predictive features. These networks have shown differences in brain activation [26], cortical thickness [25], or in connectivity [23], [27] in transgender compared with cisgender individuals. In addition to the above mentioned (i) salience and (ii) default mode networks [28], the (iii) fronto-parietal and (iv) cingulo-opercular task control networks, including dorsal and pregenual anterior cingulate cortices, are implicated in the perception of self [17]. The (v) dorsal and (vi) ventral attention networks include the temporal parietal junction and surrounding cortices important in own body perception [20]. Regions within the (vii) memory retrieval network include midline portions of the posterior cingulate shown to be important for self-perception [16]. All network-defined regions of interest (ROIs) were derived using a brain parcellation from Power et al., [29] who partitioned the brain into functional networks based on resting-state connectivity data. (See Figure S1 for node locations for a priori networks.)

For the current study, we used this knowledge of underlying biology to build a set of features capable of predicting therapeutic outcomes for GD individuals within a machine-learning framework. We focused on resting-state fMRI connectivity measures before cross-sex hormone therapy, combined with clinical data - pre-therapy body index ratings, body mass index (BMI), therapy duration, and a (simplified model) of sexual orientation (Kinsey scores) - to train and test a penalized regression model for predicting post-hormone therapy body congruence, measured by the post-hormone therapy body index scores.

## 2. Materials and Methods

### (a) Participants

Participants were recruited in Stockholm, Sweden by the Gender Team of the Center for Andrology and Sexual Medicine at Karolinska University Hospital, a center specializing in the evaluation and therapy of individuals with gender dysphoria. Adults aged 18 to 50 who were diagnosed with “Transsexualism” based on ICD-10 diagnostic criteria (F64.0, World Health Organization, 1992) (note, this term is currently outdated) or gender dysphoria (GD) based on DSM-5 [30] and sought gender-affirming medical interventions were invited to enter the study. None of the trans women or trans men had received hormonal therapy at the time of the first scanning session, or gender affirming surgery at the time of scanning sessions. Participants were excluded for previous or current hormonal therapy, any known chromosomal or hormonal disorder, or any concurrent psychiatric disorder (determined by the Mini International Neuropsychiatric Interview, MINI, Sheehan [31], neurological or other medical disorders including autism spectrum disorder, substance abuse, or the use of psychoactive medications. Participants identifying as nonbinary, or identifying other than cis- or transgender where not enrolled. All participants provided full informed consent in accordance with the Karolinska Institute ethical committee (Application # Dnr 2011/281-31/4). See Supporting Information for details of hormone therapy and gender identity.

### (b) Data Acquisition

Participants underwent an MRI scan and were evaluated with psychometric tools prior to hormonal therapy at session 1 (S1/pre-therapy). The participants were scanned and evaluated at session 2 (S2/post-therapy), on average 14 months later. We used S1 clinical measures and resting state functional connectivity (FC) as inputs to our machine learning algorithms to predict metrics of body satisfaction at S2, with the goal of determining which patients will benefit from hormone therapy - *prior to undergoing hormone therapy*.

### (c) Body Morph Task

Details of the body morph task can be found in Burke et al. 2019 [26]. Each participant was dressed in a tight, full-body unitard to provide an accurate representation of their body shape without the discomfort of being nude. Hands, feet, and head were cropped from the photos, and both front and side views were taken. Each participant’s picture was morphed towards those pictures of five different female-presenting and five different male-presenting individuals at degree intervals of 20%, using FantaMorph Software, version 5.0 (Abrosoft http://www.fantamorph.com/). Eleven morph conditions resulted, ranging between-100% morphed completely to a picture of a cisgender person with opposite birth-assigned sex to +100% morphed completely towards a picture of a cisgender person with same birth-assigned sex as the participant. Thus, 0% referred to the original unmorphed own-body image of the participant. A set of 62 images (using a randomized order and number of repetitions of the body image morphs and unmorphed own-body image) were presented for two different viewing conditions (short duration=0.5s and long duration=2s), totaling 128 trials. These images were presented using MATLAB 2012a, on a laptop computer. Each trial consisted of the image (presented for either 0.5 or 2s) followed by a 1s response screen with button press options, followed by a fixation cross. Participants were instructed to respond as quickly as possible to the question “To what degree is this picture you?” on a 4-point scale (1: 0-25% “me”, 2: 25-50% “me”, 3: 50-75% “me”, and 4: 75-100% “me”). Before the task, participants engaged in a practice session to ensure task comprehension. The clinical data was used to extract the body index (BI) [23] that was subsequently employed as a predictive feature in our machine learning algorithms.

### (d) Demographics and Psychometrics

Clinical metrics collected at S1 were used as features or covariates of non-interest: Kinsey [32] sexual orientation score, body mass index (BMI), age, therapy duration (in months from initiation of cross-hormone therapy) and birth-assigned sex. The predicted clinical measure was the body index score, calculated from the body morph task described below (Figure 1). The body morph task data were collected on a laptop, prior to the resting state MRI acquisition.

**Figure 1.**
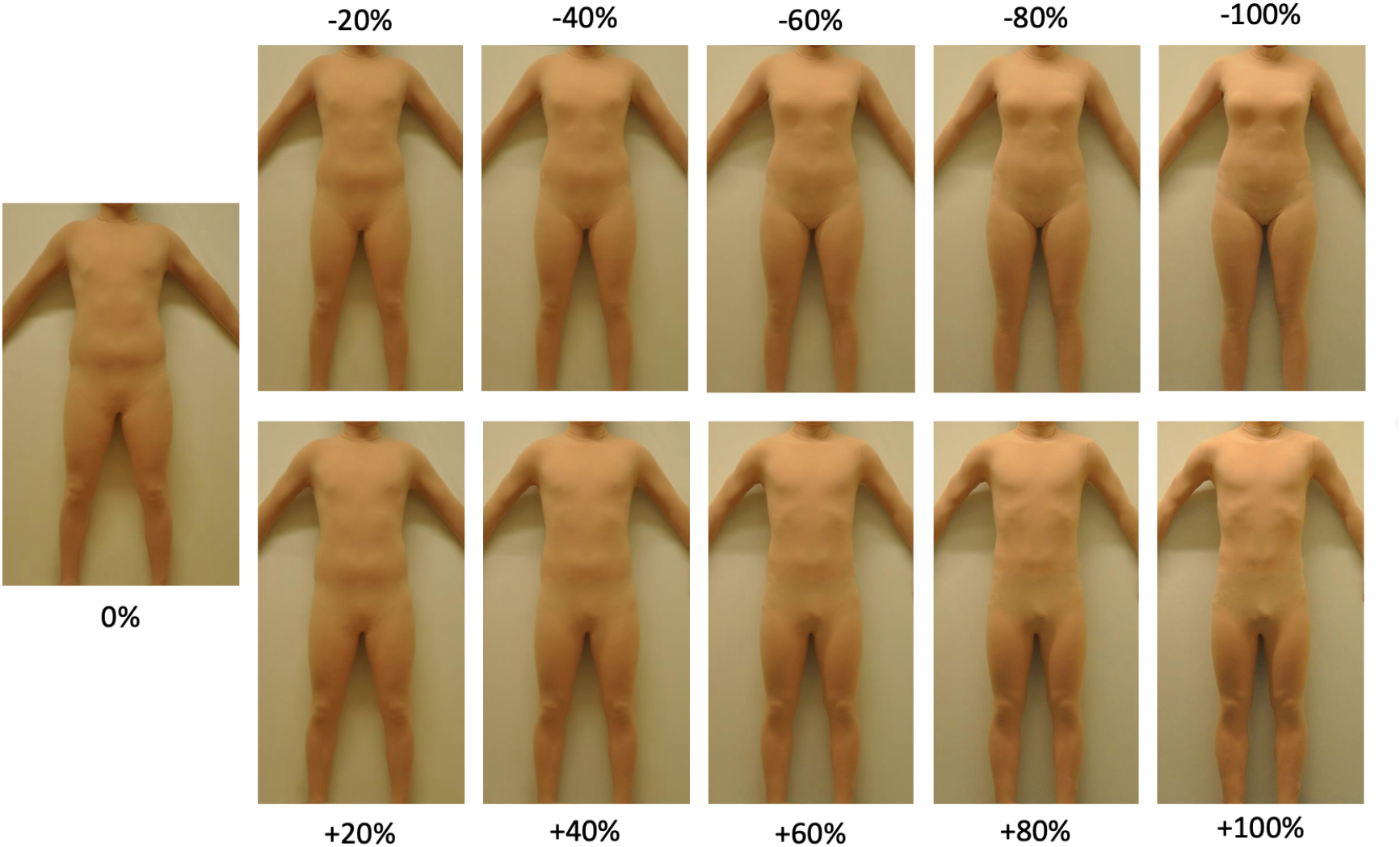
The body morph task asks subjects to rate morphed and unmorphed own-body images. Shown are examples of a front view photograph of a participant who was assigned male sex at birth morphed by 20, 40, 60, 80, and 100% to a photograph of a female (top), and male (bottom) sex-assigned-at-birth cisgender individual. Morphing to same and opposite sex-assigned-at-birth photographs are denoted (arbitrarily) by positive and negative morph degrees, respectively. Note that 100% photographs were unaltered images of another person. The 0% image is the unaltered, unmorphed own-body photograph of the participant.

To calculate the body index, we first multiplied each degree (1-4) of “self” rated for each morph with the degree of each morph. These weighted values were averaged for each participant across ratings for all images and then divided by the number of rated images, providing an average index of self-perception for each participant, weighted by how close or far from the actual self-photograph the image was morphed, and in which direction. *Positive values of the body index represent ratings toward birth-assigned sex (incongruent), while negative values represent ratings toward gender (congruent)*.

### (e) MR Data Acquisition

MRI data was acquired on a 3 Tesla MRI scanner (Discovery 3T GE-MR750, General Electric, Milwaukee, Wisconsin) using a 32-channel head coil. Resting-state functional MRI data were acquired with a gradient echo pulse sequence with: voxel size of 2.25 × 2.25 × 3 mm, TR/TE=2500/30ms, FOV=28.8cm, 45 interleaved axial slices, 90° flip angle. Each resting-state scan totaled 7min 35 s and participants were instructed to rest with eyes closed, to remain as still as possible, and not to sleep while the sequence was acquired. Structural data, 3D T1-weighted Spoiled Gradient Echo pulse sequence, were acquired with : voxel size 0.94 × 0.94 × 1 mm, TR/TE=7.91/3.06ms, TI=450ms, FOV=24 cm, 176 axial slices, 12° flip angle.

### (f) Data Analysis

MRI analysis was performed using FEAT (fMRI Expert Analysis Tool) version 5.0.8, part of FSL (FMRIB Software Library http://www.fmrib.ox.ac.uk/fsl, [33]. Bold sequences were motion-corrected (FMRIB linear image registration tool MCFLIRT), without spatial smoothing, and individual participants’ resting state data were denoised using FSL’s AROMA tool (non-aggressive denoising option). Functional images were registered to their respective T1-weighted images (FMRIB non-linear image registration tool, FNIRT) after brain extraction using FSL’s BET, and then to the MNI-152 brain for functional connectivity analysis. Two participants were excluded from further analyses due to head motion greater than 1.5 mm.

### (g) ROI Selection

We used a functionally-defined set of ROIs (10mm diameter spheres) that have been previously mapped to functional networks [29], [34]. We narrowed our scope to seven *a priori* networks that covered regions and networks with functional and/or structural differences between cisgender and GD or transgender individuals: default mode, fronto-parietal, dorsal attention, salience, cingulo-opercular, memory retrieval, and ventral attention. This resulted in 264 ROIs, each of which was tagged with one of seven functional-network identities.

### (h) ROI correlation matrices

We collected resting-state data from each participant prior to cross-hormone therapy and used the denoised images to determine connectivity among the ROIs. For each participant we computed the mean BOLD activity within each of the ROIs at every time point (every 2.5 seconds), resulting in a time course of mean ROI activity. We then computed a pairwise Pearson correlation matrix for each mean time course resulting in 264 × 264 matrices containing the pairwise functional connectivity values (r-values) across all ROIs.

### (i) Feature Creation

We indexed each correlation matrix depending on network identity, extracting the lower diagonal of each matrix and then identifying the rows and columns corresponding to the functional connectivity across ROIs within a specific network. These constituted the functional connectivity (FC) feature sets. We included these feature sets plus clinical data, as the “hand-selected” features that we tested in the machine learning algorithms. See Figure 2 for a flow chart of the machine learning analysis. Unless otherwise indicated, all predictions were tested for significance of p<.05 and were subjected to the Bonferroni method to correct for multiple comparisons. The Bonferroni-adjusted p-value, p_bf_ <0.006 (p<0.05/9), was adjusted by the seven networks in our a priori hypotheses, and two combinations of networks (all seven networks and the fronto-parietal and cingulo-opercular networks together).

**Figure 2.**
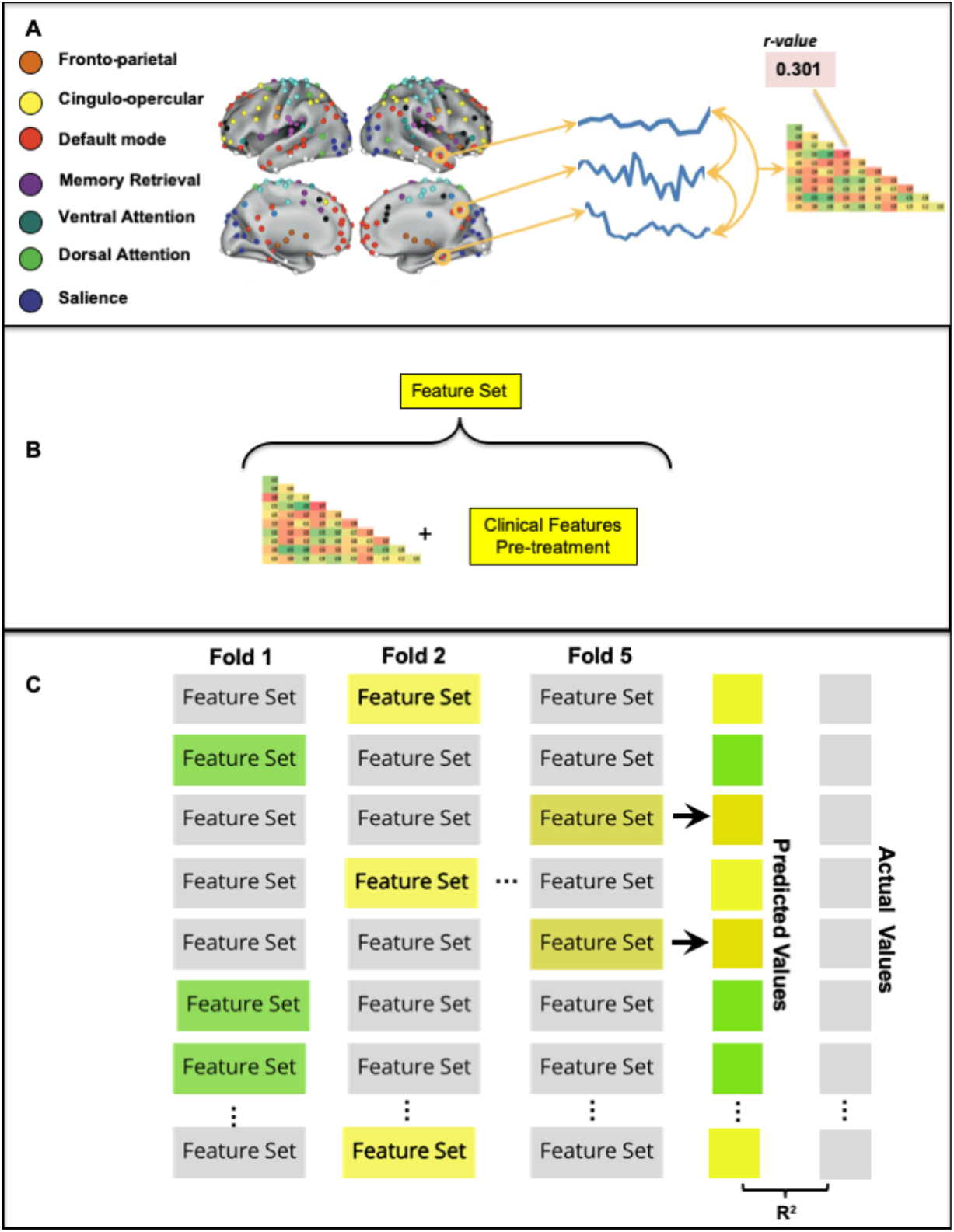
Analysis flow chart. (A) The average resting-state activity within ROIs from seven functional brain networks defined by Power (30) was used to create a mean BOLD time course. Pairwise Pearson correlations of these time courses resulted in a functional connectivity matrix specific for each network. (B) The lower diagonal of each participant’s network-specific matrix was concatenated with the participant’s pre-therapy clinical features scores to create a feature set for that participant. (C) The LASSO regression model was trained on n−5 participants’ feature sets and their associated post-therapy body index scores and used to predict each of the left-out participant’s post-therapy body index scores. Left-out participants are denoted as highlighted feature sets (only three shown here). This process was repeated until all participants had been left out in a fold of the cross-validation and had been assigned a predicted post-therapy body index score. We correlated the array of predicted values (ŷ) with the actual values (y), resulting in Pearson’s R^2^, a measure of our model’s feature-dependent ability to capture the outcome variable variance across participants. Note that due to our participant sample size (n = 25), one fold of the cross-validation left out five participants.

### (j) Machine learning regression analysis predicting post-therapy body index using absolute shrinkage and selection operator (LASSO)

We built a LASSO [35],[34] regression model on n-5 (20%) participants using their feature sets. LASSO was chosen as the regression model of choice due to its ability to handle large feature sets, impose a self-directed feature selection, and output continuous variables. Using each trained model’s intercept term and beta coefficients, we calculated a predicted measure of interest from each left-out participant’s feature set. After obtaining a prediction for each participant (ŷ), we correlated the array of predicted values with the actual values (y) to quantify the model’s feature-dependent ability to capture the variance in clinical measures across participants.

Clinical features included pre-therapy body index rating, therapy duration, BMI, and Kinsey scores. Age was treated as a covariate of non-interest and iteratively regressed out of each feature in the feature set prior to classification. We did not include birth-assigned sex as a covariate, as it is considered within the body index ratings. For predicting body index for short duration trials, we used the pre-therapy body index ratings for short duration trials as a feature, likewise, for predicting pre-therapy body index for long duration trials, we used the body index ratings for long duration trials as a feature. In a preliminary analysis, we examined the effect of therapy duration in our analyses by including therapy duration either as a covariate or as a clinical feature. For six of the seven networks, using therapy duration as a feature was more predictive than using therapy duration as a covariate (Table S1); therefore, results reported for body index are for therapy duration as a feature.

### (k) Machine learning prediction of post-therapy body index using ridge regression

To provide a robustness check on our LASSO predictions, we re-ran the above machine learning analysis using ridge [36] regression. Ridge regression is appropriate when the predictor features potentially have collinearity. (See Table S2, also see Table S3 for additional post-hoc tests of specificity.)

## 3. Results

### (a) Participants

Twenty-five adults ages 18-50, mean years 25.2 (SD 7.8), with DSM-diagnosed GD participated in the study. Data from 16 women, assigned male at birth, and 9 men, assigned female at birth, were combined for all analyses. See Table 1 for demographics.

**Table 1.** Demographics, clinical values and ratings of the participants (N=25).

| Characteristic | Value | SD | P value<br>T-test | P value<br>correlation |
| --- | --- | --- | --- | --- |
| Trans women/Trans men | 16/9 |  |  |  |
| Age | 25.2 | 7.8 |  |  |
| BMI | 24.1 | 5.5 |  |  |
| Kinsey scores | 4.0 | 2.0 |  |  |
| Years of education | 13.3 | 1.9 |  |  |
| Therapy duration (months) | 14.3 | 5.4 |  |  |
| Body index pre-therapy<br>short duration trials | -10.4 | 21.8 |  |  |
| Body index post-therapy<br>short duration trials | -23.1 | 25.7 | p=0.002* | p<0.001* |
| Body index pre-therapy<br>long duration trials | -11.1 | 33.5 |  |  |
| Body index post-therapy<br>long duration trials | -21.3 | 31.7 | p=0.040* | p<0.001* |

\* Paired one-tailed, t-test, comparing pre- versus post-hormone therapy.
\* Correlation, comparing pre- versus post-hormone therapy.
Negative values of the body index represent (arbitrarily) ratings toward pictures of opposite sex-assigned-at-birth individuals (congruent with sense of self in those with gender dysphoria).

### (b) Body Congruence Changes

For short duration trials, body index scores changed in the direction of increased congruence pre- to post hormone-therapy in 18 of 25 participants (72%), see Figure S2. The pre-therapy mean was -10.4 (SD, 21.8); the post-therapy mean was -23.1 (SD, 25.7), t_24_ = 3.1, p = 0.002, 1-tailed. Similarly, for long duration trials, change in the direction of increased congruence was observed in 15 of 25 participants (60%). The pre-therapy mean was -11.1 (SD, 33.5); the post-therapy mean was - 21.3 (SD, 31.7), t_24_ = 1.8, p =0.04, 1-tailed (Table 2). More-negative scores on the body index are indicative of greater congruence of their body with their gender identity. There were associations in a comparison of pre- and post-therapy values of body index scores: body index ratings for short duration trials (R^2^ = 0.41, p = 0.006) and body index ratings for long duration trials (R^2^ = 0.39, p = 0.008). There was a trend for an association between therapy duration and body index ratings after hormone therapy for short duration trials, (R^2^ = 0.11, p = 0.11), see Table S4.

**Table 2.**
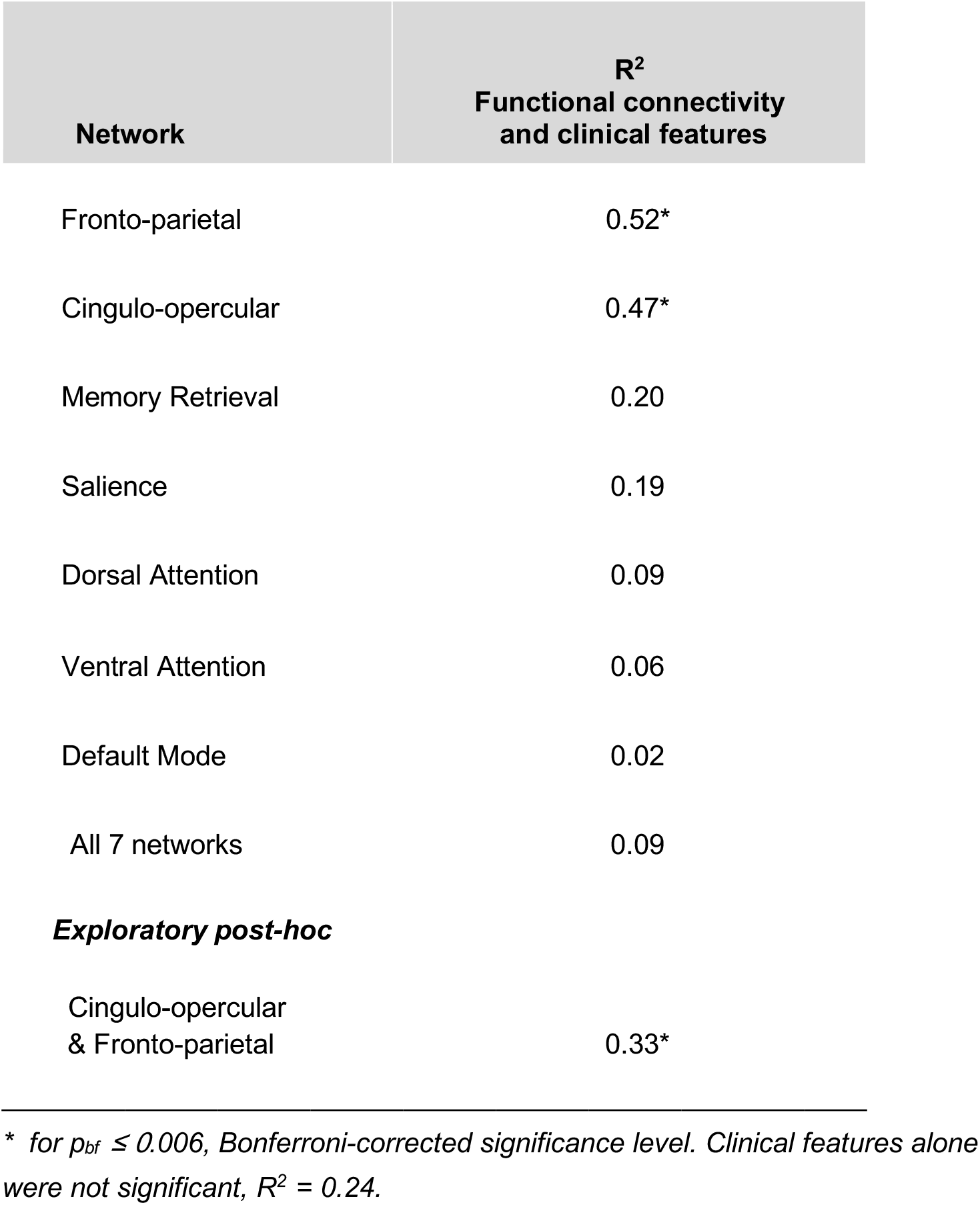
Associations between predicted post-therapy body congruence for seven brain functional connectivity networks, combined with clinical features, using multivariate analysis.

| Network | R<sup>2</sup><br>Functional connectivity<br>and clinical features |
| --- | --- |
| Fronto-parietal | 0.52* |
| Cingulo-opercular | 0.47* |
| Memory Retrieval | 0.20 |
| Salience | 0.19 |
| Dorsal Attention | 0.09 |
| Ventral Attention | 0.06 |
| Default Mode | 0.02 |
| All 7 networks | 0.09 |
| **Exploratory post-hoc** |  |
| Cingulo-opercular<br>& Fronto-parietal | 0.33* |

\* for $p_{bf} \leq 0.006$ , Bonferroni-corrected significance level. Clinical features alone were not significant, $R^2 = 0.24$ .

### (c) Body congruence prediction

#### Predicting post-therapy body index ratings using LASSO machine learning regression

For these analyses, we used functional connectivity (FC) from resting-state fMRI data either a) across all nodes composing an individual network or b) across all aggregate nodes composing multiple networks. Short-duration trials (Table 2), but not long-duration trials (Table S5), predicted post-therapy body congruence. Clinical data points that were also leveraged alongside the FC values in the feature set included pre-therapy body index ratings, sexual orientation (Kinsey scores), BMI, and time from initiation of therapy. See Table S6 for similar results, without considering therapy duration.

##### Individual a priori networks

The associations between the algorithm’s predicted post-therapy body index ratings (ŷ) and the actual ratings (y) was statistically significant when using two of the seven a priori networks, the fronto-parietal network and the cingulo-opercular network, alongside the clinical features. When only the clinical features were considered in the model, there was not an association between predicted (ŷ) and actual (y) body index values, R^2^ = 0.24, p = 0.013; however, when the model included the clinical features and the functional connectivity, there were associations for the fronto-parietal network, R^2^ = 0.52, p < 0.001 and for the cingulo-opercular network, R^2^ = 0.47, p < 0.001, (Figure 3, Table 2), for Bonferroni corrected, p_bf_< 0.006.

**Figure 3.**
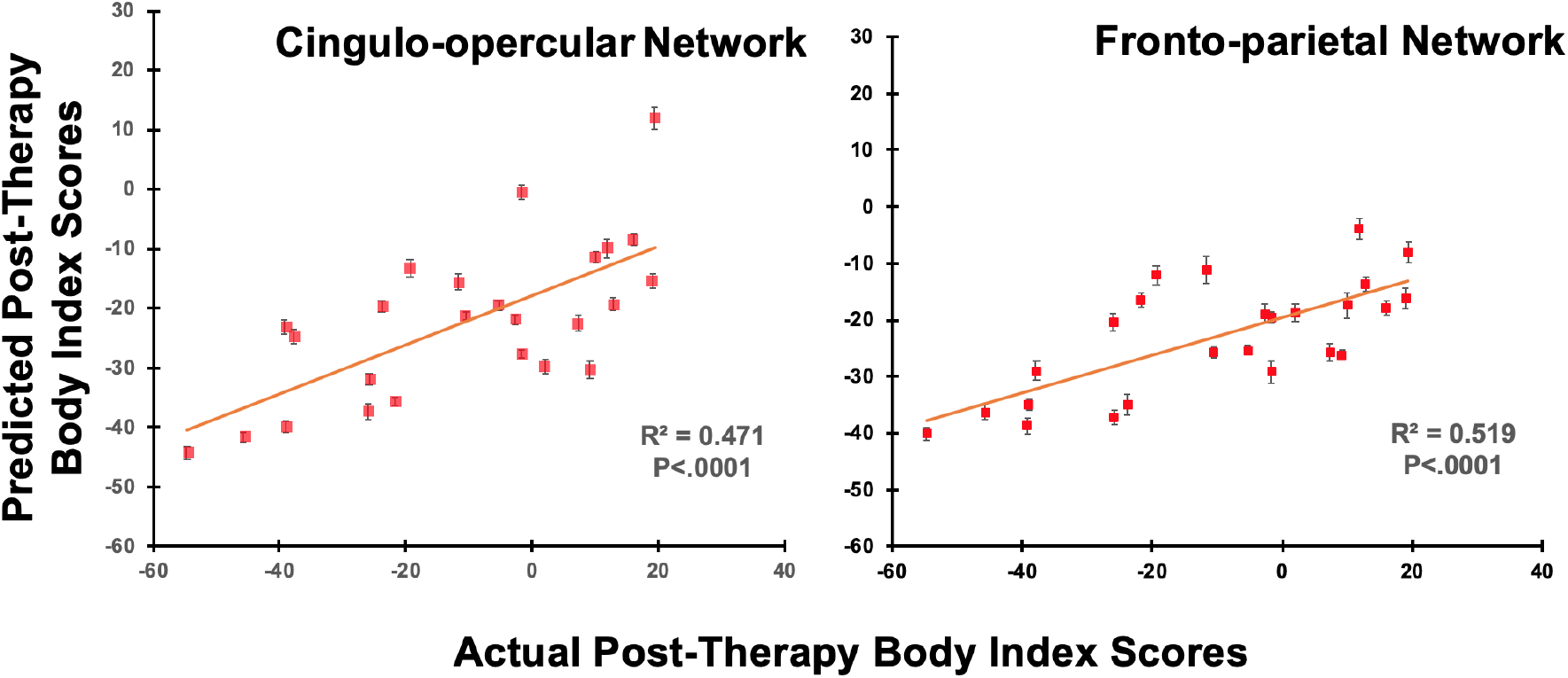
Associations between the distributions of body index predictions and actual post-therapy values are shown in scatter plots. These LASSO cross-validation models used feature sets that included pre-therapy functional connectivity from the cingulo-opercular network (Left) and the fronto-parietal network (Right), in addition to clinical features. Error bars are standard-errors across the 100 cross-validation predictions for each individual. The Bonferroni-corrected significance level is p_bf_ ≤ 0.006.

##### Aggregated a priori networks

Including all seven of the a priori networks in the model along with pre-therapy clinical features resulted in a lower value of R^2^ = 0.09, p = 0.149. However, combining the two networks (cingulo-opercular and fronto-parietal) that showed significant associations between predicted and actual body index ratings above, with clinical features, resulted in an association of R^2^ = 0.32, p = 0.003.

##### Predicting post-therapy body index ratings using ridge machine learning regression

We additionally tested predictions using ridge regression. The results when including the same clinical and network features were similar to those using the LASSO models, showing associations for cingulo-opercular R^2^ = 0.41, p = 0.001, and fronto-parietal networks R^2^ = 0.32, p = 0.003, and combining the two networks R^2^ = 0.32, p = 0.003; all significant for p_bf_ < 0.006, Table S2. Clinical features alone were not associated, R^2^ = 0.25, p = 0.010.

##### Post Hoc prediction of pre-therapy body index ratings

The association between the *predicted pre-therapy* body index ratings and *actual pre-therapy* ratings for short duration trials was not significant when using clinical features and the FC of any of our a priori networks.

## 4. Discussion

This study in individuals with gender dysphoria tested whether multivariate pattern recognition using neurobiological features from resting state brain connectivity along with clinical features could be used to predict therapeutic response to cross-hormone therapy. The goal was to predict, on an individual basis, the important clinical outcome of body congruence in those with gender dysphoria after hormone therapy by using brain functional connectivity data from a short (7.5 minute) MRI scan, BMI, and body congruence ratings before hormone therapy. Multivariate connectivity in the cingulo-opercular and fronto-parietal networks before hormone therapy explained a high proportion of the variance in individual body congruence after hormone therapy. Clinical variables alone *were not able to explain* body congruence using the body index ratings. These findings have implications for identifying those who will benefit more or less from hormone therapy. Furthermore, these results support our previous finding using anatomical metrics [25,28], and contribute to identifying the specific brain networks in GD, *prior to therapy*, whose connectivity patterns are critical with respect to hormone therapy effects.

The predictive model that we built and tested was able to explain 52% of the variance in body congruence subsequent to cross-hormone therapy. The predictive power of multivariate connectivity was substantiated by the overlapping results of LASSO and ridge machine learning algorithms, which converged to provide evidence that functional connectivity from cingulo-opercular and fronto-parietal networks can be used prior to initiation of hormone therapy to predict body congruence after hormone therapy. Exploiting multivariate techniques may thus provide additional insight into not only the neurobiological bases but also the sociological, cultural, social, and psychological bases of gender and body satisfaction. A recent study [37] employed machine learning, based on functional connectivity, to predict, better than chance, self-reported gender identity in four groups (trans/cis, women/men). Further, including therapy duration in our analyses generates an algorithm that has beta weights for this as a feature. Therefore, if a new person came into a clinic, entering a specific future time point after therapy initiation, e.g. 6 mo, or 1 year, could provide an estimate of that person’s body congruence at that time point. The algorithm that did not include therapy duration was similarly predictive (Table S6).

This work can be conceived of in the context of the evolving concept of using a functional connectome as a “fingerprint” that is an index of highly individualized latent neural organization [38]. This latent neural organization is linked to response tendencies, processing of stimuli, multisensory integration, and patterns of conscious and unconscious thinking [38]. The most predictive networks in the current study, the fronto-parietal and cingulo-opercular networks, comprise important regions implicated in self-identity, self-referential thinking, as well as supporting top-down control of executive functioning [39]. While the fronto-parietal and cingulo-opercular networks are largely intraconnected and separable, they also appear to communicate, or perhaps compete, for control functions [40]. Koush et al [41] found that the superior frontal gyrus (within the fronto-parietal network) modulates self-referential processes in the temporal parietal junction as well as affective valuation in ventromedial prefrontal cortex - which in turn is an important hub of the default mode network. Because in the current study the fronto-parietal and cingulo-opercular networks were predictive of post-therapy but not pre-therapy body congruence, these networks may be more specifically involved in cognitive reorganization that may occur with hormone therapy. Perhaps this could involve the directed control of conscious perception of body and body changes as they fit into one’s sense of gender self-identity. If so, the pre-therapy connectivity in these networks may be markers of the degree that individuals’ brain network organization is able to update one’s sense of self as one’s body and hormonal milieu changes. As these are cognitive control networks it might point to the directed control of self-referential thought processes with body self-awareness. This is potentially informative of the neurological underpinnings of gender identity in relation to body and hormonal status among transgender individuals as they transition.

The observation that these networks that significantly predicted p*ost-therapy body congruence* were not also associated with *pre-therapy body congruence* suggests that these networks may be more specifically involved in cognitive reorganization occurring with hormone therapy. One speculation, for example, is that this may identify those individuals whose multivariate connectivity pattern may index better or worse ability to bring their experience of their gender identity in line with the perception of their post-hormone bodies.

Connectivity before hormone therapy within the fronto-parietal and cingulo-opercular networks was most predictive of body congruence for short duration trials. It is not clear why ratings of short-duration trials were more predictive of body congruence than ratings of long-duration trials. One possibility is that the longer two-second trials allow rumination that interferes with the “truer” reflexive responses required by the half-second trials. Related to this, some of the ratings for long duration trials may have been influenced by individuals’ difficulty viewing the body images for longer times because of continued dysphoria triggered by viewing the images, in addition to longstanding patterns of avoidance of viewing their bodies, leading to ratings that may have been made in a cursory way and thereby not reflecting their true degree of congruence.

The body index has been used in other studies [15,22,24,25] as a metric of body congruence. Another scale measuring body congruence is the self-report TCS. We did not have TCS scores for most of the participants in this study so did not include that metric in this analysis; however, we have examined TCS scores in two ongoing datasets and found trends for positive associations between TCS scores and the body index in individuals with GD (Supporting Information). This, in addition to significant changes in the body index pre- to post-therapy and an association between therapy duration and changes in body index (Figure S3) lends support for the body index as a measure of therapy-sensitive body congruence.

A limitation of the current study is sample size. The cross-validation approach of leaving out 20% of the participants for model testing reduced the likelihood of overfitting. However, larger datasets would provide the opportunity to split the participants into training and testing groups for a more robust validation. In addition, validation in fully independent test sets, ideally in different settings, would determine if the results may be generalizable to other populations with GD in different geographical locations and cultural and societal environments. Further, our results are limited to binary-identifying individuals and thus cannot necessarily be generalized to nonbinary and differently gendered populations. Importantly, the use of more complex models that include other variables, for example minority stress, societal attitudes, a more nuanced characterization of gender identities, and other sociological factors, would more comprehensively capture likely contributing factors to treatment outcomes, and might result in a better-performing algorithm. Due to sample size limitations we were not able to consider trans men and trans women separately in this study. Future work should do so, since a recent report [15] has shown that trans women generally had lower body index ratings than trans men for short duration trials and trans men rated images morphed opposite to their birth-assigned sex slightly higher than trans women. This is in line with other work [42] that found trans men had a more positive body image than trans women. Future investigations of the mechanisms underlying the regions within the fronto-parietal and cingulo-opercular networks that drive the results seen here are warranted. In addition, while the current investigation adds to evidence that hormone therapy may enhance body congruence in gender dysphoria, changes in gonadal steroid levels have been shown to affect mood and cognition [43], [44]. This study illustrates the potential for predicting hormone therapy responsiveness in transgender individuals with gender dysphoria. One goal of this line of research is to enhance therapy for the individual by providing an optimal therapy plan, with consideration for the physical and psychological commitment of the patient, as well as time and cost of therapy. Hormone therapy is expensive and requires years of commitment in most cases. A more immediate practical application of these results, if replicated, would be applying the algorithm proposed here to identify individuals for whom it may be more critical to apply therapies in addition to hormones - such as gender affirming surgeries to optimize body congruence. This approach may also help identify those for whom standard hormone protocols are not expected to work as well and who may need different estrogen or androgen antagonists and/or alternate types or regimens of sex hormones, or no sex hormones. Further, results from the study could help us understand what pre-therapy brain networks may be involved in post-therapy body congruence and thus establish biomarkers that could potentially be used to develop novel ways of improving body congruence. In sum, this study contributes to understanding neurophysiological aspects of therapy in gender dysphoria. Insights from this research could contribute to future therapy guidelines for gender dysphoria.

## Data Availability

Data availability is governed by the Swedish IRB.

## Ethics

The Karolinska Institute ethical committee (Application # Dnr 2011/281-31/4) approved this study. All participants gave informed consent at the beginning of the study.

## Data accessibility

Data and materials are available on the Open Science Framework.

## Authors’ Contributions

TDM, NR, IS, and JDF designed research; TDM, NR, AM, MH, IS, and JSF performed research; TDM, NR, IS, JV, and JDF contributed new analytic tools; TDM analyzed data; and TDM, NR, MH, AM, IS, JV and JDF wrote the paper. All authors gave final approval for publication.

## Competing interests

The authors declare no competing interests.

## Funding

This work was supported by the Swedish Science Council (Dnr 200 7-3107 to I.S) and the National Institutes of Health Eunice Kennedy Shriver National Institute of Child Health and Human Development (1R01HD087712 to I.S. and J.F.). J.V. was supported by the Vocatio Foundation (Belgium).

## Acknowledgements

We thank Natalie Rotstein for assistance with data validation.

## Notes

### Competing Interest Statement

The authors have declared no competing interest.

### Author Declarations

All participants provided full informed consent in accordance with the Karolinska Institute ethical committee (Application # Dnr 2011/281-31/4).

